# Healthcare workers hospitalized due to COVID-19 have no higher risk of death than general population. Data from the Spanish SEMI-COVID-19 Registry

**DOI:** 10.1101/2020.11.23.20236810

**Authors:** Jesús Díez-Manglano, Marta Nataya Solís Marquínez, Andrea Álvarez García, Nicolás Alcalá-Rivera, Irene Maderuelo Riesco, Martín Gericó Aseguinolaza, José Luis Beato Pérez, Manuel Mendez Bailon, Ane Elbire Labirua-Iturburu Ruiz, Miriam García Gómez, Carmen Martinez Cilleros, Paula Maria Pesqueira Fontan, Lucy Abella Vázquez, Julio César Blázquez Encinar, Ramon Boixeda, Ricardo Gil Sánchez, Andrés de la Peña Fernández, Jose Loureiro Amigo, Joaquin Escobar Sevilla, Marcos Guzmán Garcia, María Dolores Martín Escalante, Jeffrey Oskar Magallanes Gamboa, Angel Luís Martínez Gonzalez, Carlos Lumbreras Bermejo, Juan Miguel Antón Santos, for the SEMI-COVID-19 Network

**Affiliations:** Internal Medicine Department, Royo Villanova Hospital, Zaragoza, Spain; Internal Medicine Department, San Agustin University Hospital, Avilés (Asturias), Spain; Internal Medicine Department, Albacete University Hospital, Albacete, Spain; Internal Medicine Department, San Carlos Clinical Hospital, Madrid, Spain; Internal Medicine Department, Santa Marina Hospital, Bilbao, Spain; Internal Medicine Department, Urduliz Alfredo Espinosa Hospital, Urdúliz (Vizcaya), Spain; Internal Medicine Department, HLA Moncloa Hospital, Madrid, Spain; Internal Medicine Department, Santiago Clinical Hospital, Santiago de Compostela (A Coruña), Spain; Internal Medicine Department, Ntra Sra Candelaria University Hospital, Santa Cruz de Tenerife, Spain; Internal Medicine Department, Torrevieja University Hospital, Torrevieja (Alicante), Spain; Internal Medicine Department, Mataró Hospital, Mataró (Barcelona), Spain; Internal Medicine Department, La Fe University Hospital, Valencia, Spain; Internal Medicine Department, Son Llàtzer University Hospital, Palma de Mallorca, Spain; Internal Medicine Department, Moisès Broggi Hospital, Sant Joan Despí (Barcelona), Spain; Internal Medicine Department, Virgen de las Nieves University Hospital, Granada, Spain; Internal Medicine Department, San Juan de la Cruz Hospital, Úbeda (Jaén), Spain; Internal Medicine Department, Costa del Sol Hospital, Marbella (Málaga), Spain; Internal Medicine Department, Nuestra Señora del Prado Hospital, Talavera de la Reina (Toledo), Spain; Internal Medicine Department, León University Hospital, León, Spain; Internal Medicine Department, 12 de Octubre University Hospital, Madrid, Spain; Internal Medicine Department, Infanta Cristina University Hospital, Parla (Madrid), Spain

**Keywords:** SARS-CoV-2, coronavirus, COVID-19, Spain, healthcare workers, mortality

## Abstract

**Aim:** To determine whether healthcare workers (HCW) hospitalized in Spain due to COVID-19 have a worse prognosis than non-healthcare workers (NHCW).

**Methods:** Observational cohort study based on the SEMI-COVID-19 Registry, a nationwide registry that collects sociodemographic, clinical, laboratory, and treatment data on patients hospitalised with COVID-19 in Spain. Patients aged 20-65 years were selected. A multivariate logistic regression model was performed to identify factors associated with mortality.

**Results:** As of 22 May 2020, 4393 patients were included, of whom 419 (9.5%) were HCW. Median (interquartile range) age of HCW was 52 (15) years and 62.4% were women. Prevalence of comorbidities and severe radiological findings upon admission were less frequent in HCW. There were no difference in need of respiratory support and admission to intensive care unit, but occurrence of sepsis and in-hospital mortality was lower in HCW (1.7% vs. 3.9%; p=0.024 and 0.7% vs. 4.8%; p<0.001 respectively). Age, male sex and comorbidity, were independently associated with higher in-hospital mortality and healthcare working with lower mortality (OR 0.219, 95%CI 0.069-0.693, p=0.01). 30-days survival was higher in HCW (0.968 vs. 0.851 p<0.001).

**Conclusions:** Hospitalized COVID-19 HCW had fewer comorbidities and a better prognosis than NHCW. Our results suggest that professional exposure to COVID-19 in HCW does not carry more clinical severity nor mortality.

## INTRODUCTION

As of 30 October 2020, coronavirus disease 2019 (COVID-19), caused by severe acute respiratory syndrome coronavirus 2 (SARS-CoV-2), has affected 44,592,789 people worldwide (1). Spain has been one of the countries with the highest number of confirmed cases and deaths.

Healthcare workers (HCW) are at high risk of infection with SARS-CoV-2 because of their exposure to infected patients. Several seroepidemiological studies have shown discordant results. In New York and Sweden the seroprevalence of SARS-CoV-2 among hospital HCW was high compared with the community (2,3). However, in Germany the seroprevalence in HCW was very low, 0.33% (4). A recent systematic review and meta-analysis, that included 11 studies, reported that the overall proportion of HCW who were SARS-CoV-2 positive among all COVID-19 patients was 10.1% (5). In Spain, 20.4% of confirmed COVID-19 cases were HCW (6).

In Scotland, HCW and their households contributed a sixth of COVID-19 admissions to hospital (7). In Spain and USA, 10% and 8% of HCW with COVID-19 were hospitalized respectively (6,8). Young women nurses were more frequently infected (9-11). Comorbidities were frequent in HCW hospitalized with COVID-19, particularly diabetes, hypertension, obesity, asthma and immunodepression (6,10,11).

There is still controversy over the risk of death in HCW with COVID-19. While it is high in Mexico, it is low in Germany and Malaysia (12,13). The main objectives of this study were to describe the clinical characteristics and outcomes of HCW hospitalised in Spain due to SARS-CoV-2 infection, and to determine if working in healthcare is associated with higher rates of complications and mortality.

## METHODS

### Study design and population

The SEMI-COVID-19 Registry is an ongoing, nationwide, multicentre, observational retrospective registry, participated by 150 hospital centres throughout Spain. Detailed features of the registry have been reported elsewhere (14-16). A total of 10,600 consecutive patients were recruited from March 1, 2020 to May 22, 2020.

### Inclusion criteria

The SEMI-COVID-19 Registry includes patients > 18 years admitted to hospital with COVID-19 confirmed microbiologically by reverse transcription polymerase chain reaction (RT-PCR) testing of a nasopharyngeal swab sample, sputum specimen or bronchoalveolar lavage. The exclusion criteria were subsequent admission of the same patient or denial or withdrawal of informed consent. This study analyses the subpopulation of patients between 20 and 65 years of age. In Spain, 20 years is the youngest possible age for working in healthcare and 65 years is the retirement age. HCW were defined as physicians, nurses, nurse aides, and non-healthcare professionals such as the administrative and cleaning staff of hospitals and healthcare centres.

### Procedures and variables

Admission and treatment of patients took place at the discretion of the attending physicians based on their clinical judgment, local protocols, and the updated recommendations of the Spanish Ministry of Health.

Data were collected retrospectively in an online electronic data capture system and were extracted from electronic health records. Approximately 300 variables were collected including epidemiological data, RT-PCR data, personal medical and medication history, symptoms and physical examinations findings at admission, laboratory and diagnostic imaging tests, pharmacological treatment and ventilator support during hospitalization, complications and death during hospitalization, and readmissions and survival 30 days after diagnosis. Comorbidity was assessed using the Charlson Comorbidity Index (CCI) (17). Degree of independence was classified as independent or mild, moderate, and severe. Obesity was defined as body mass index >30 kg/m^2^.

The main endpoint was mortality during admission. Intensive care unit (ICU) admission, days in the ICU, invasive or non-invasive ventilation, all-cause re-admission, 30-days all-cause mortality, and length-of-stay were secondary endpoints.

### Ethical aspects

The study was carried out in accordance with the Declaration of Helsinki. The processing of personal data strictly complied with Spanish Law 14/2007, of July 3, on Biomedical Research and Spanish Organic Law 3/2018, of 5 December, on the Protection of Personal Data and the Guarantee of Digital Rights.The study was approved by the Provincial Research Ethics Committee of Málaga (Spain) following the recommendation of the Spanish Agency of Medicines and Medical Products (AEMPS, for its initials in Spanish). All patients—or their caregivers, in the event they presented with cognitive impairment—gave their informed oral consent.

### Statistical analysis

The patients were divided into two groups: HCW and non HCW (NHCW). Continuous variables were tested for normal distribution with the Kolmogorov-Smirnov test. Quantitative variables are expressed as mean (standard deviation, SD) or median [interquartile range]. Comparisons between groups were made using Student’s t-test and the Mann-Whitney U test. Categorical variables are expressed as absolute frequencies and percentages. Comparisons between them were made using the chi-square test with the Yates correction and with the Fisher’s exact test when necessary.

Two multivariate logistic regression models were performed to analyse the association between working in healthcare and mortality,. The first model included age, sex, ethnicity, CCI score, and healthcare working. The second model included the previous variables and added the comorbidities with a statistical significance p<0.1 in the univariate model.

In all cases, statistical significance was established as p<0.05. Statistical analysis was carried out using Statistical Package for the Social Sciences (SPSS) 21.0 software for Windows.

## RESULTS

**Figure 1** shows the flowchart for patient inclusion. A total of 4,393 patients were included, of which 419 (9.5%) were HCW. Among HCW, 142 (33.9%) were medical doctors, 107 (25.5%) were nurses, 98 (23.4%) were nurse aides, and 72 (17.2%) held other positions within healthcare. The departments that most infected patients worked in were primary care (16.6%), the emergency department (11.3%), and internal medicine (11.3%).

**Figure 1.**
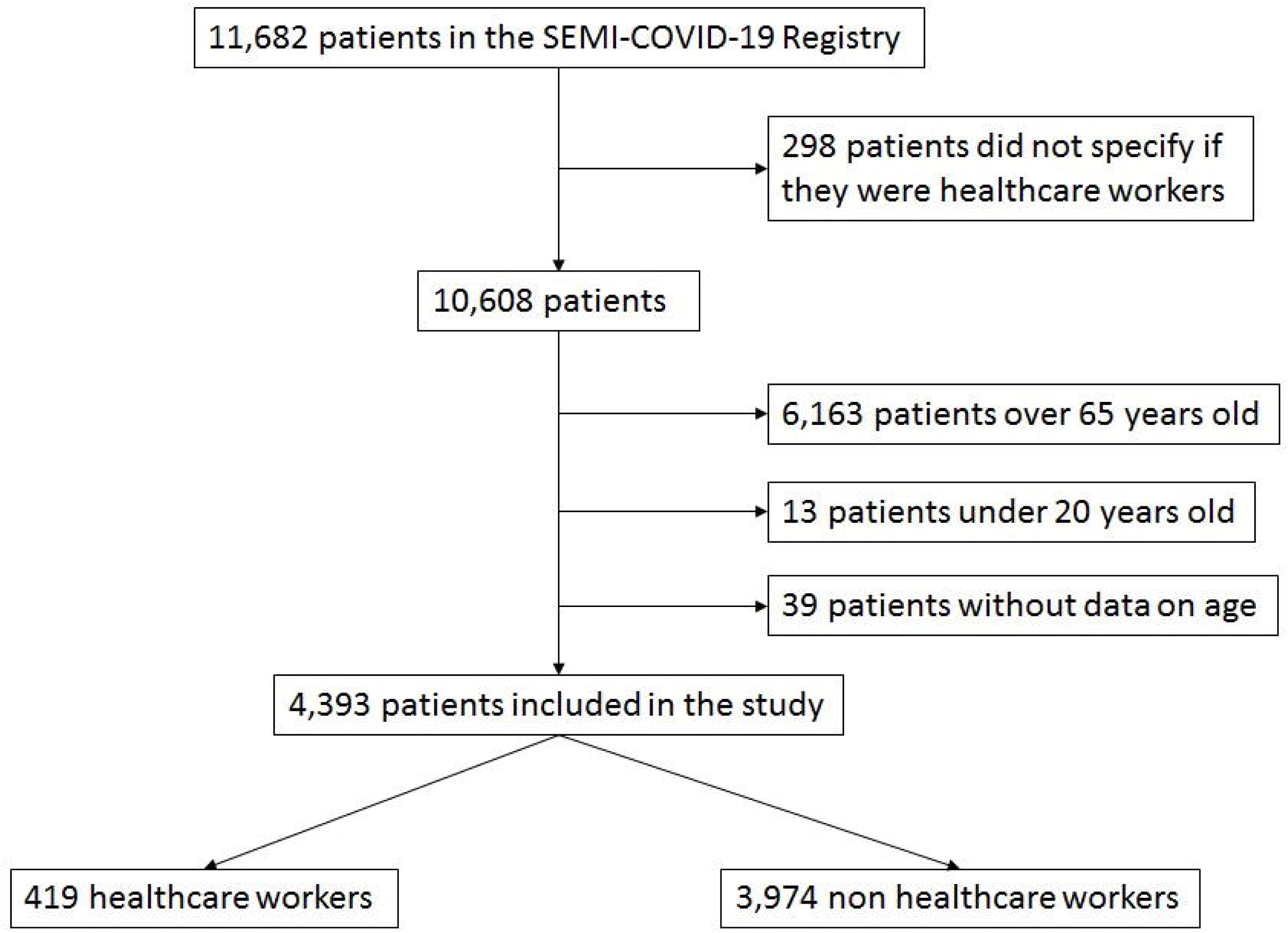
Patient inclusion flowchart.

### Demographic and clinical characteristics

Baseline demographic and clinical characteristics and comorbidities are shown in **Table 1**. HCW were more often Caucasian women, and reported more frequent contact with a COVID-19 patient (57.8% vs. 22.1%, p<0.001). Moderate and severe dependence was more frequent in NHCW. There was no difference in comorbidity measured as Charlson index score, but the prevalence of comorbidities as alcohol use disorder, hypertension, dyslipidemia, obesity, diabetes, myocardial infarction, stroke, dementia, chronic obstructive pulmonary disease, obstructive sleep apnea-hypopnea syndrome, chronic kidney disease and malignancy was higher in NHCW.

**Table 1.**
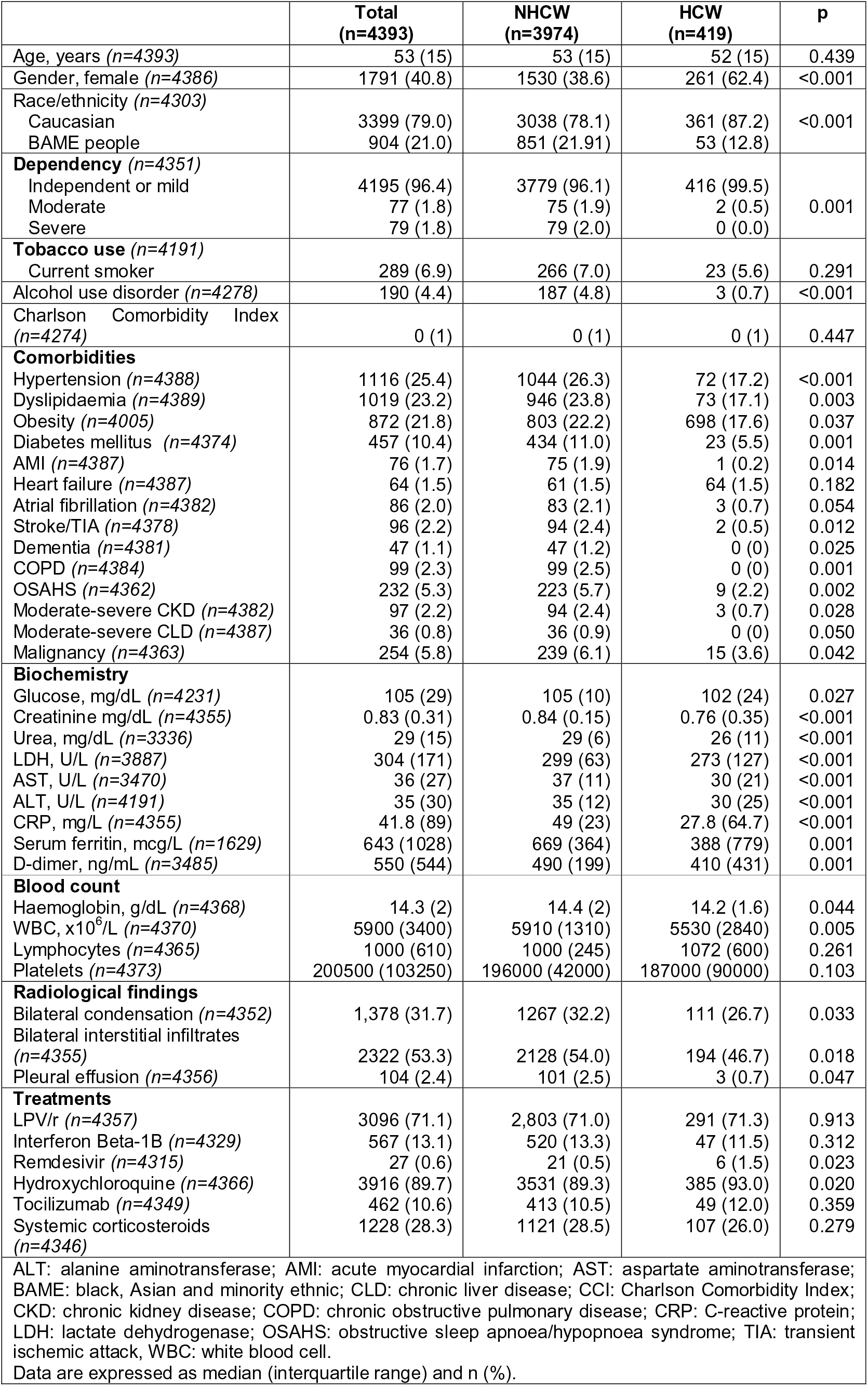
Sociodemographic characteristics, comorbidities and treatment of patients included.

|  | **Total**<br>**(n=4393)** | **NHCW**<br>**(n=3974)** | **HCW**<br>**(n=419)** | **p** |
| --- | --- | --- | --- | --- |
| Age, years (n=4393) | 53 (15) | 53 (15) | 52 (15) | 0.439 |
| Gender, female (n=4386) | 1791 (40.8) | 1530 (38.6) | 261 (62.4) | <0.001 |
| Race/ethnicity (n=4303) |  |  |  |  |
| Caucasian | 3399 (79.0) | 3038 (78.1) | 361 (87.2) | <0.001 |
| BAME people | 904 (21.0) | 851 (21.91) | 53 (12.8) |  |
| **Dependency (n=4351)** |  |  |  |  |
| Independent or mild | 4195 (96.4) | 3779 (96.1) | 416 (99.5) | 0.001 |
| Moderate | 77 (1.8) | 75 (1.9) | 2 (0.5) |  |
| Severe | 79 (1.8) | 79 (2.0) | 0 (0.0) |  |
| **Tobacco use (n=4191)** |  |  |  |  |
| Current smoker | 289 (6.9) | 266 (7.0) | 23 (5.6) | 0.291 |
| Alcohol use disorder (n=4278) | 190 (4.4) | 187 (4.8) | 3 (0.7) | <0.001 |
| Charlson Comorbidity Index (n=4274) | 0 (1) | 0 (1) | 0 (1) | 0.447 |
| **Comorbidities** |  |  |  |  |
| Hypertension (n=4388) | 1116 (25.4) | 1044 (26.3) | 72 (17.2) | <0.001 |
| Dyslipidaemia (n=4389) | 1019 (23.2) | 946 (23.8) | 73 (17.1) | 0.003 |
| Obesity (n=4005) | 872 (21.8) | 803 (22.2) | 698 (17.6) | 0.037 |
| Diabetes mellitus (n=4374) | 457 (10.4) | 434 (11.0) | 23 (5.5) | 0.001 |
| AMI (n=4387) | 76 (1.7) | 75 (1.9) | 1 (0.2) | 0.014 |
| Heart failure (n=4387) | 64 (1.5) | 61 (1.5) | 64 (1.5) | 0.182 |
| Atrial fibrillation (n=4382) | 86 (2.0) | 83 (2.1) | 3 (0.7) | 0.054 |
| Stroke/TIA (n=4378) | 96 (2.2) | 94 (2.4) | 2 (0.5) | 0.012 |
| Dementia (n=4381) | 47 (1.1) | 47 (1.2) | 0 (0) | 0.025 |
| COPD (n=4384) | 99 (2.3) | 99 (2.5) | 0 (0) | 0.001 |
| OSAHS (n=4362) | 232 (5.3) | 223 (5.7) | 9 (2.2) | 0.002 |
| Moderate-severe CKD (n=4382) | 97 (2.2) | 94 (2.4) | 3 (0.7) | 0.028 |
| Moderate-severe CLD (n=4387) | 36 (0.8) | 36 (0.9) | 0 (0) | 0.050 |
| Malignancy (n=4363) | 254 (5.8) | 239 (6.1) | 15 (3.6) | 0.042 |
| **Biochemistry** |  |  |  |  |
| Glucose, mg/dL (n=4231) | 105 (29) | 105 (10) | 102 (24) | 0.027 |
| Creatinine mg/dL (n=4355) | 0.83 (0.31) | 0.84 (0.15) | 0.76 (0.35) | <0.001 |
| Urea, mg/dL (n=3336) | 29 (15) | 29 (6) | 26 (11) | <0.001 |
| LDH, U/L (n=3887) | 304 (171) | 299 (63) | 273 (127) | <0.001 |
| AST, U/L (n=3470) | 36 (27) | 37 (11) | 30 (21) | <0.001 |
| ALT, U/L (n=4191) | 35 (30) | 35 (12) | 30 (25) | <0.001 |
| CRP, mg/L (n=4355) | 41.8 (89) | 49 (23) | 27.8 (64.7) | <0.001 |
| Serum ferritin, mcg/L (n=1629) | 643 (1028) | 669 (364) | 388 (779) | 0.001 |
| D-dimer, ng/mL (n=3485) | 550 (544) | 490 (199) | 410 (431) | 0.001 |
| **Blood count** |  |  |  |  |
| Haemoglobin, g/dL (n=4368) | 14.3 (2) | 14.4 (2) | 14.2 (1.6) | 0.044 |
| WBC, x10<sup>6</sup>/L (n=4370) | 5900 (3400) | 5910 (1310) | 5530 (2840) | 0.005 |
| Lymphocytes (n=4365) | 1000 (610) | 1000 (245) | 1072 (600) | 0.261 |
| Platelets (n=4373) | 200500 (103250) | 196000 (42000) | 187000 (90000) | 0.103 |
| **Radiological findings** |  |  |  |  |
| Bilateral condensation (n=4352) | 1,378 (31.7) | 1267 (32.2) | 111 (26.7) | 0.033 |
| Bilateral interstitial infiltrates (n=4355) | 2322 (53.3) | 2128 (54.0) | 194 (46.7) | 0.018 |
| Pleural effusion (n=4356) | 104 (2.4) | 101 (2.5) | 3 (0.7) | 0.047 |
| **Treatments** |  |  |  |  |
| LPV/r (n=4357) | 3096 (71.1) | 2,803 (71.0) | 291 (71.3) | 0.913 |
| Interferon Beta-1B (n=4329) | 567 (13.1) | 520 (13.3) | 47 (11.5) | 0.312 |
| Remdesivir (n=4315) | 27 (0.6) | 21 (0.5) | 6 (1.5) | 0.023 |
| Hydroxychloroquine (n=4366) | 3916 (89.7) | 3531 (89.3) | 385 (93.0) | 0.020 |
| Tocilizumab (n=4349) | 462 (10.6) | 413 (10.5) | 49 (12.0) | 0.359 |
| Systemic corticosteroids (n=4346) | 1228 (28.3) | 1121 (28.5) | 107 (26.0) | 0.279 |

ALT: alanine aminotransferase; AMI: acute myocardial infarction; AST: aspartate aminotransferase; BAME: black, Asian and minority ethnic; CLD: chronic liver disease; CCI: Charlson Comorbidity Index; CKD: chronic kidney disease; COPD: chronic obstructive pulmonary disease; CRP: C-reactive protein; LDH: lactate dehydrogenase; OSAHS: obstructive sleep apnoea/hypopnoea syndrome; TIA: transient
ischemic attack, WBC: white blood cell. Data are expressed as median (interquartile range) and n (%).

The median time from first symptoms to admission was 7 [5-9] days, without difference between HCW and NHCW. There were some differences in symptoms and physical examination findings. Dry cough (72.3% vs. 67.3%, p=0.003), asthenia (54.6% vs. 44.8%, p<0.001), arthralgia (48.1% vs. 39.2%, p<0.001), ageusia (14.6% vs. 9.4%, p=0.001) and anosmia (14.6% s. 8.7%, p<0.001) were more frequent in HCW, and temperature ≥ 38°C (74.4% vs. 68,5%, p=0.007) and oxygen saturation ≤ 92% (24.2% vs 11.8%, p<0.001) in NHCW.

At admission the levels of serum glucose, creatinine, urea, lactate dehydrogenase, aspartate aminotransferase, alanine aminotransferase, C-reactive protein, ferritin, D-dimer, hemoglobin and the count of white blood cells were lower in HCW. NHCW presented more frequently severe radiological findings—i.e. pleural effusion, bilateral condensation, and bilateral interstitial infiltrates (all p≤0.025).

### Treatments

There were no differences in the treatment for COVID-19 disease between HCW and NHCW except for hydroxychloroquine and remdesivir (93% vs. 89.3%; p=0.02 and 1.5% vs. 0.5%; p=0.0.23 respectively).

### Outcomes

Sepsis was more frequent in NHCW (3.9% vs. 1.7%; p=0.024). There were no differences in the occurrence of other complications, the need of respiratory support or ICU admission. The length of hospital stay was 8 (7) days without difference among HCW and NHCW. During hospitalization 194 (4.4%) patients died. In-hospital mortality was lower in HCW (0.7% vs 4.8%; p<0.001). The readmission rate was 2.9%. Half of readmissions were due to COVID-19 disease (**Table 2**). The 30-days survival was 96.8% in HCW and 85.1% in NHCW (p=0.001). **Figure 2** shows the Kaplan-Meier survival curve.

**Table 2.** Outcomes.

|  | Total | NHCW | HCW | p |
| --- | --- | --- | --- | --- |
| **Complications** |  |  |  |  |
| Bacterial pneumonia ($n=4356$) | 330 (7.6) | 305 (7.7) | 25 (6.0) | 0.209 |
| ARDS ($n=4355$) | 1001 (23.0) | 919 (23.3) | 82 (19.8) | 0.101 |
| Acute kidney failure ($n=4353$) | 243 (5.6) | 228 (5.8) | 15 (3.6) | 0.068 |
| Sepsis ($n=4351$) | 160 (3.7) | 153 (3.9) | 7 (1.7) | 0.024 |
| Shock ($n=4349$) | 132 (3.0) | 125 (3.2) | 7 (1.7) | 0.094 |
| Thromboembolic disease ($n=4350$) | 71 (1.6) | 64 (1.6) | 7 (1.7) | 0.921 |
| **Respiratory support** |  |  |  |  |
| High flow nasal cannula ($n=4334$) | 327 (7.5) | 293 (7.5) | 34 (8.2) | 0.578 |
| Noninvasive mechanical ventilation ($n=4357$) | 174 (4.0) | 156 (4.0) | 18 (4.3) | 0.707 |
| Invasive mechanical ventilation ($n=4357$) | 314 (7.2) | 291 (7.4) | 23 (5.5) | 0.168 |
| **Intensive care unit (ICU)** |  |  |  |  |
| Admission to ICU ($n=4385$) | 415 (9.5) | 371 (9.4) | 44 (10.5) | 0.435 |
| Days in the ICU | 11 (10) | 11 (11) | 8.5 (10) | 0.099 |
| **Death and readmission** |  |  |  |  |
| Hospital length-of-stay, days ($n=4392$) | 8 (7) | 8 (8) | 7 (7) | 0.067 |
| In-hospital death ($n=4393$) | 194 (4.4) | 191 (4.8) | 3 (0.7) | $<0.001$ |
| Readmission ($n=4194$) | 121 (2.9) | 112 (2.9) | 9 (2.3) | 0.449 |
| ARDS: acute respiratory distress syndrome; ICU: intensive care unit<br>Data are expressed as n (%) and median [interquartile range] |  |  |  |  |

**Figure 2.**
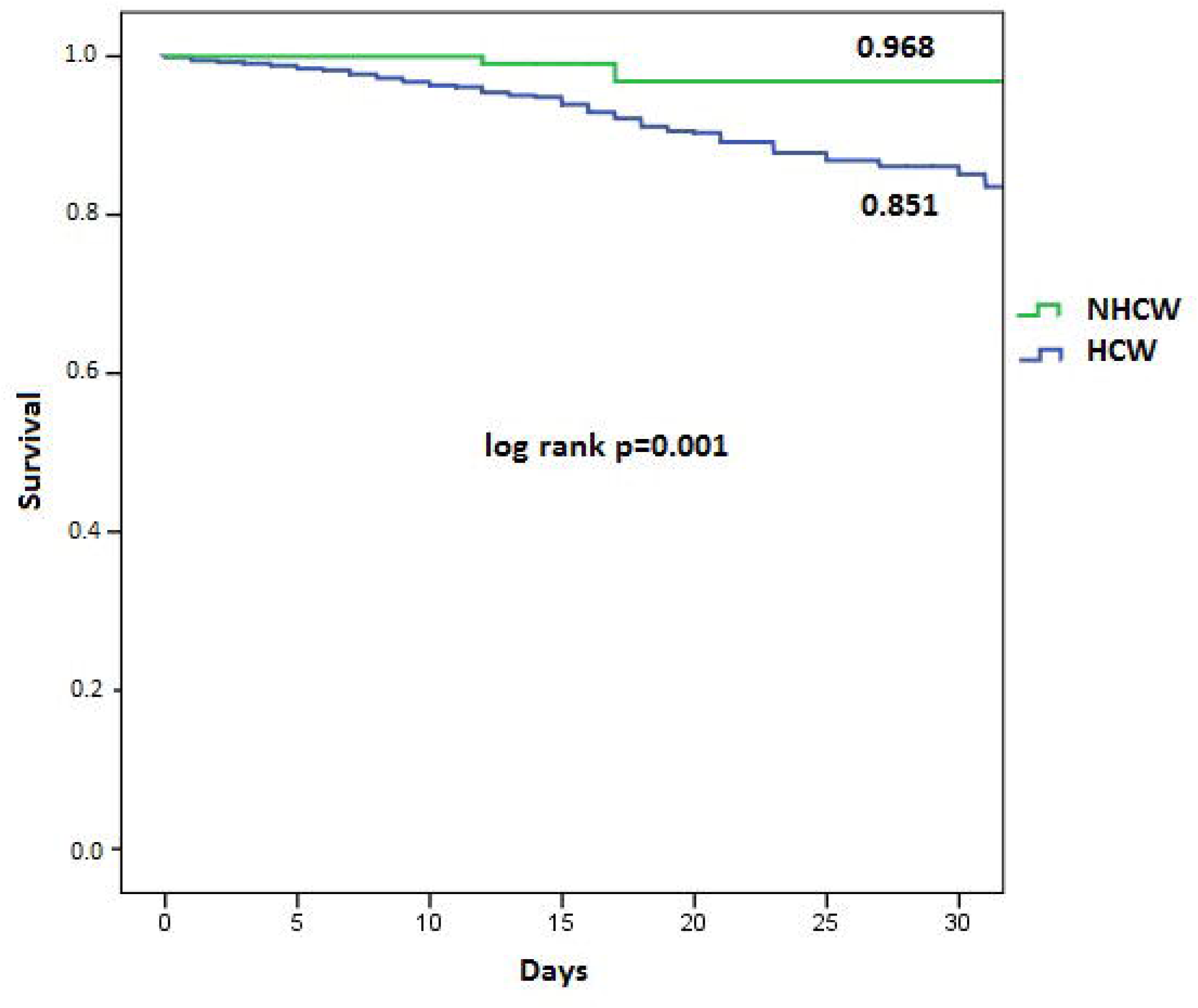
30-days Kaplan-Meier survival curves.

The factors associated with in-hospital mortality are shown in **Table 3**. In the first multivariate analysis model, age, male sex and Charlson Comorbidity index score were associated with higher in-hospital mortality and healthcare working with lower mortality (OR 0.219, 95%CI 0.069-0.693, p=0.01). In the second model, including comorbidities, healthcare working was also associated with a lower in-hospital mortality (OR 0.285 95%CI 0.089-0.908; p=0.034).

**Table 3.** Factors associated with mortality.

|  | Univariate |  | Multivariate |  |
| --- | --- | --- | --- | --- |
| Variable | OR (95%CI) | p | OR (95%CI) | p |
| **Model 1** |  |  |  |  |
| Age | 1.073 (1.053-1.095) | <0.001 | 1.370 (1.287-1.458) | <0.001 |
| Male sex | 1.884 (1.366-2.599) | <0.001 | 1.062 (1.040-1.084) | <0.001 |
| BAME | 0.743 (0.505-1.094) | 0.132 |  |  |
| HCW | 0.143 (0.045-0.449) | 0.001 | 0.219 (0.069-0.693) | 0.01 |
| CCI | 1.423 (1.340-1.512) | <0.001 | 2.088 (1.474-2.957) | <0.001 |
| **Model 2** |  |  |  |  |
| Age | 1.073 (1.053-1.095) | <0.001 | 1.055 (1.031-1.081) | <0.001 |
| Male sex | 1.884 (1.366-2.599) | <0.001 | 1.712 (1.159-2.530) | 0.007 |
| BAME | 0.743 (0.505-1.094) | 0.132 |  |  |
| HCW | 0.143 (0.045-0.449) | 0.001 | 0.285 (0.089-0.908) | 0.034 |
| Alcohol | 2.642 (1.604-4.351) | <0.001 | 1.266 (0.674-2.379) | 0.463 |
| Smoking | 2.491 (1.625-3.820) | <0.001 | 2.436 (1.473-4.030) | 0.001 |
| Hypertension | 2.362 (1.762-3.166) | <0.001 | 1.360 (0.923-2.004) | 0.120 |
| Dyslipidemia | 1.886 (1.393-2.552) | <0.001 | 0.819 (0.543-1.236) | 0.342 |
| Obesity | 2.029 (1.471-2.798) | <0.001 | 1.679 (1.136-2.481) | 0.009 |
| Diabetes | 2.870 (2.030-4.059) | <0.001 | 1.464 (0.926-2.316) | 0.103 |
| Acute myocardial infarction | 2.609 /1.236-5.508) | 0.012 | 1.278 (0.518-3.156) | 0.595 |
| Heart failure | 3.660 (1.782-7.519) | <0.001 | 1.158 (0.426-3.147) | 0.773 |
| Atrial fibrillation | 2.279 (1.085-4.788) | 0.03 | 0.939 (0.376-2.344) | 0.893 |
| Stroke/TIA | 3.913 (2.178-7.033) | <0.001 | 0.952 (0.417-2.171) | 0.907 |
| Dementia | 8.773 (4.551-16.910) | <0.001 | 8.884 (3.800-20.772) | <0.001 |
| COPD | 6.017 (3.600-10.057) | <0.001 | 2.330 (1.233-4.403) | 0.009 |
| OSAHS | 3.165 (2.058-4.867) | <0.001 | 1.659 (0.955-2.884) | 0.072 |
| Moderate-severe CKD | 5.351 (3.138-9.126) | <0.001 | 3.649 (1.889-7.051) | <0.001 |
| Moderate-severe CLD | 5.375 (2.324-12.429) | <0.001 | 1.845 (0.657-5.180) | 0.245 |
| Malignancy | 3.710 (2.490-5.527) | <0.001 | 3.058 (1.891-4.943) | <0.001 |
| BAME: black, Asian and minority ethnic; CCI: Charlson Comorbidity Index score; CI: confidence interval; CKD: chronic kidney disease; CLD; chronic liver disease; COPD: chronic obstructive pulmonary disease; HCW: healthcare workers; OR: odds ratio; OSAHS: obstructive sleep apnoea/hypopnoea syndrome; TIA: transient ischemic attack |  |  |  |  |

## DISCUSSION

The main findings of our study were that hospitalised HCW had less severe COVID-19 and lower mortality.

The demographic characteristics of our patients were consistent with other reports (14,18,19). Worldwide, men were more likely to be infected by SARS-CoV-2 than women. However, among HCW, women were the most affected (8,9,13,20). We think that this difference is due to the higher proportion of females in healthcare professions. When we compared our HCW cohort with those reported in other studies, our patients were more than ten years older (8-10,13,20,21). This could be due to the different inclusion criteria used in each study. In the USA, Hughes et al (8) reported on the characteristics of 100,570 HCW with a median age of 41 years, of whom only 6832 were hospitalised. Wang et al included 80 HCW hospitalised in Wuhan, of whom 57 were confirmed cases and 23 were clinical diagnosis (11). In our cohort, only hospitalised patients between 20 and 65 years old with a confirmed diagnosis of COVID-19 were included, while the rest included all HCW with COVID-19.

An interesting finding in our study was that upon admission HCW presented milder symptoms, such as loss of smell or taste and arthralgia, less severe radiological findings and lower lactate dehydrogenase, C-reactive protein, serum ferritin and D-dimer levels. And all of it even though there was no difference between HCW and NHCW in time from onset of symptoms and admission. There was not an explanation for this, but we hypothesize that it could be due to HCW were hospitalised earlier and more easily than NHCW.

ARDS is overwhelmingly the main cause of death in hospitalised COVID-19 patients. In our study, sepsis was less frequent in HCW but there was no difference in ARDS, the rest of complications, the need of respiratory support nor the ICU admission. In-hospital and 30-days mortality were lower in HCW. In a systematic review Similar results were reported in the systematic review by Sahu et al. (5). A healthy worker effect could explain these results. Severily ill and chronically disabled are ordinarily excluded from employment (22). The difference observed in prevalence of comorbidities between HCW and NHCW supports this explanation. The better clinical and analytical profile of the HCW at admission may be due to their knowledge of mild symptoms of COVID-19 and their ability to identify them in themselves. In this regard, increased education on the earliest and mildest symptoms of COVID-19 could help NHCW to recognize and to report them to a healthcare centre earlier in the course of their disease.

There are geographical differences in mortality observed in hospitalised COVID-19 HCW. In a teaching hospital in Belgium the mortality of HCW was 0.5% (20). In a single-centre study in Wuhan, in-hospital death in HCW with confirmed SARS-CoV-2 infection was 1.7%, more than twice in our study (11). The mortality in a multicentre study in New York City was 21%, far higher than that we observed (19). Also, high mortality, 14.7%, was reported in Brazil (10). In Mexico, a mortality of 2% of HCW with COVID-19 was reported (21). Their mortality was higher than ours even though only 9% of them needed to be hospitalized. The authors explain their high mortality because of different reasons. On the one hand, in Mexico there is a high prevalence of comorbidities which are associated with severe COVID-19. On the other hand, due to structural inequalities, their healthcare system is highly heterogeneous and there is a remarkable amount of marginalized communities. Therefore, the prevalence of comorbidities, the level of economic wealth, and the organization of healthcare in the different countries could explain these differences observed in the mortality.

Several studies have reported that age, male sex and comorbidity were associated with higher mortality in COVID-19 patients (23-26). However the research about healthcare working as risk factor of mortality is scarce. HCW worry and are afraid to be infected and die for COVID-19. Our results confirm that COVID-19 is less severe and leads to less mortality in HCW. This is one of the novel contributions of our study.

Among the strengths of our study are its multicentre design, the inclusion of patients from the entire country, and the large number of patients included, which provides an adequate statistical power to confirm hypotheses. However, our study also has limitations. Only hospitalised patients were included, so it is not possible to extrapolate our results to non-hospitalised patients. The large number of researchers involved and variability in the availability of data in each hospital could have led to information bias. Finally, the voluntary participation of each centre could have caused selection bias.

In conclusion, HCW had fewer comorbidities, milder symptoms, and a better prognosis than the NHCW. Our results suggest that professional exposure to COVID-19 in HCW does not lead to greater clinical severity nor mortality.

## Data Availability

We used data of the SEMI-COVID-19-Registry, which has been previously described (Casas JM. Rev Clin Esp 2020; 220: 480-94). Due to legal issues, some restrictions apply to our data and they cannot be uploaded to any public server. However, we can share the data as a request to the Principal Investigator of the EpiChron Cohort Study at.

## Acknowledgments

**List of the SEMI-COVID-19 Network members**

**Coordinator of the SEMI-COVID-19 Registry:** José Manuel Casas Rojo.

**SEMI-COVID-19 Scientific Committee Members:** José Manuel Casas Rojo, José Manuel Ramos Rincón, Carlos Lumbreras Bermejo, Jesús Millán Núñez-Cortés, Juan Miguel Antón Santos, Ricardo Gómez Huelgas.

**SEMI-COVID-19 Registry Coordinating Center:** S & H Medical Science Service.

**Members of the SEMI-COVID-19 Group**

<u>H. U. 12 de Octubre. Madrid</u>

Paloma Agudo de Blas, Coral Arévalo Cañas, Blanca Ayuso, José Bascuñana Morejón, Samara Campos Escudero, María Carnevali Frías, Santiago Cossio Tejido, Borja de Miguel Campo, Carmen Díaz Pedroche, Raquel Diaz Simon, Ana García Reyne, Lucia Jorge Huerta, Antonio Lalueza Blanco, Jaime Laureiro Gonzalo, Carlos Lumbreras Bermejo, Guillermo Maestro de la Calle, Barbara Otero Perpiña, Diana Paredes Ruiz, Marcos Sánchez Fernández, Javier Tejada Montes.

<u>H. U. Gregorio Marañón. Madrid</u>

Laura Abarca Casas, Álvaro Alejandre de Oña, Rubén Alonso Beato, Leyre Alonso Gonzalo, Jaime Alonso Muñoz, Crhistian Mario Amodeo Oblitas, Cristina Ausín García, Marta Bacete Cebrián, Jesús Baltasar Corral, Maria Barrientos Guerrero, Alejandro Bendala Estrada, María Calderón Moreno, Paula Carrascosa Fernández, Raquel Carrillo, Sabela Castañeda Pérez, Eva Cervilla Muñoz, Agustín Diego Chacón Moreno, Maria Carmen Cuenca Carvajal, Sergio de Santos, Andrés Enríquez Gómez, Eduardo Fernández Carracedo, María Mercedes Ferreiro-Mazón Jenaro, Francisco Galeano Valle, Alejandra Garcia, Irene Garcia Fernandez-Bravo, María Eugenia García Leoni, Maria Gomez Antunez, Candela González San Narciso, Anthony Alexander Gurjian, Lorena Jiménez Ibáñez, Cristina Lavilla Olleros, Cristina Llamazares Mendo, Sara Luis García, Víctor Mato Jimeno, Clara Millán Nohales, Jesús Millán Núñez-Cortés, Sergio Moragón Ledesma, Antonio Muiño Miguez, Cecilia Muñoz Delgado, Lucía Ordieres Ortega, Susana Pardo Sánchez, Alejandro Parra Virto, María Teresa Pérez Sanz, Blanca Pinilla Llorente, Sandra Piqueras Ruiz, Guillermo Soria Fernández-Llamazares, María Toledano Macías, Neera Toledo Samaniego, Ana Torres do Rego, Maria Victoria Villalba Garcia, Gracia Villarreal, María Zurita Etayo.

<u>Hospital Universitari de Bellvitge. L’Hospitalet de Llobregat</u>

Xavier Corbella, Abelardo Montero, Jose María Mora-Luján.

<u>C. H. U. de Albacete. Albacete</u>

Jose Luis Beato Pérez, Maria Lourdes Sáez Méndez.

<u>H. U. La Paz-Cantoblanco-Carlos III. Madrid</u>

Jorge Álvarez Troncoso, Francisco Arnalich Fernández, Francisco Blanco Quintana, Carmen Busca Arenzana, Sergio Carrasco Molina, Aranzazu Castellano Candalija, Germán Daroca Bengoa, Alejandro de Gea Grela, Alicia de Lorenzo Hernández, Alejandro Díez Vidal, Carmen Fernández Capitán, Maria Francisca García Iglesias, Borja González Muñoz, Carmen Rosario Herrero Gil, Juan María Herrero Martínez, Víctor Hontañón, Maria Jesús Jaras Hernández, Carlos Lahoz, Cristina Marcelo Calvo, Juan Carlos Martín Gutiérrez, Monica Martinez Prieto, Elena Martínez Robles, Araceli Menéndez Saldaña, Alberto Moreno Fernández, Jose Maria Mostaza Prieto, Ana Noblejas Mozo, Carlos Manuel Oñoro López, Esmeralda Palmier Peláez, Marina Palomar Pampyn, Maria Angustias Quesada Simón, Juan Carlos Ramos Ramos, Luis Ramos Ruperto, Aquilino Sánchez Purificación, Teresa Sancho Bueso, Raquel Sorriguieta Torre, Clara Itziar Soto Abanedes, Yeray Untoria Tabares, Marta Varas Mayoral, Julia Vásquez Manau.

<u>Complejo Asistencial de Segovia. Segovia</u>

Eva María Ferreira Pasos, Daniel Monge Monge, Alba Varela García.

<u>H. U. Puerta de Hierro. Majadahonda</u>

María Álvarez Bello, Ane Andrés Eisenhofer, Ana Arias Milla, Isolina Baños Pérez, Javier Bilbao Garay, Silvia Blanco Alonso, Jorge Calderón Parra, Alejandro Callejas Díaz, José María Camino Salvador, M^<u>a</u>^ Cruz Carreño Hernández, Valentín Cuervas-Mons Martínez, Sara de la Fuente Moral, Miguel del Pino Jimenez, Alberto Díaz de Santiago, Itziar Diego Yagüe, Ignacio Donate Velasco, Ana María Duca, Pedro Durán del Campo, Gabriela Escudero López, Esther Expósito Palomo, Ana Fernández Cruz, Esther Fiz Benito, Andrea Fraile López, Amy Galán Gómez, Sonia García Prieto, Claudia García Rodríguez-Maimón, Miguel Ángel García Viejo, Javier Gómez Irusta, Edith Vanessa Gutiérrez Abreu, Isabel Gutiérrez Martín, Ángela Gutiérrez Rojas, Andrea Gutiérrez Villanueva, Jesús Herráiz Jiménez, Pedro Laguna del Estal, M^<u>a</u>^ Carmen Máinez Sáiz, Cristina Martín Martín, María Martínez Urbistondo, Fernando Martínez Vera, Susana Mellor Pita, Patricia Mills Sánchez, Esther Montero Hernández, Alberto Mora Vargas, Cristina Moreno López, Alfonso Ángel-Moreno Maroto, Victor Moreno-Torres Concha, Ignacio Morrás De La Torre, Elena Múñez Rubio, Ana Muñoz Gómez, Rosa Muñoz de Benito, Alejandro Muñoz Serrano, Jose María Palau Fayós, Ilduara Pintos Pascual, Antonio Ramos Martínez, Isabel Redondo Cánovas del Castillo, Alberto Roldán Montaud, Lucía Romero Imaz, Yolanda Romero Pizarro, Mónica Sánchez Santiuste, David Sánchez Órtiz, Enrique Sánchez Chica, Patricia Serrano de la Fuente, Pablo Tutor de Ureta, Ángela Valencia Alijo, Mercedes Valentín-Pastrana Aguilar, Juan Antonio Vargas Núñez, Jose Manuel Vázquez Comendador, Gema Vázquez Contreras, Carmen Vizoso Gálvez.

<u>H. Miguel Servet. Zaragoza</u>

Gonzalo Acebes Repiso, Uxua Asín Samper, María Aranzazu Caudevilla Martínez, José Miguel García Bruñén, Rosa García Fenoll, Jesús Javier González Igual, Laura Letona Giménez, Mónica Llorente Barrio, Luis Sáez Comet.

<u>H. U. La Princesa. Madrid</u>

María Aguilera García, Ester Alonso Monge, Jesús Álvarez Rodríguez, Claudia Alvarez Varela, Miquel Berniz Gòdia, Marta Briega Molina, Marta Bustamante Vega, Jose Curbelo, Alicia de las Heras Moreno, Ignacio Descalzo Godoy, Alexia Constanza Espiño Alvarez, Ignacio Fernández Martín-Caro, Alejandra Franquet López-Mosteiro, Gonzalo Galvez Marquez, María J. García Blanco, Yaiza García del Álamo Hernández, Clara García-Rayo Encina, Noemí Gilabert González, Carolina Guillamo Rodríguez, Nicolás Labrador San Martín, Manuel Molina Báez, Carmen Muñoz Delgado, Pedro Parra Caballero, Javier Pérez Serrano, Laura Rabes Rodríguez, Pablo Rodríguez Cortés, Carlos Rodriguez Franco, Emilia Roy-Vallejo, Monica Rueda Vega, Aresio Sancha Lloret, Beatriz Sánchez Moreno, Marta Sanz Alba, Jorge Serrano Ballester, Alba Somovilla, Carmen Suarez Fernández, Macarena Vargas Tirado, Almudena Villa Marti.

<u>H. U. de A Coruña. A Coruña</u>

Alicia Alonso Álvarez, Olaya Alonso Juarros, Ariadna Arévalo López, Carmen Casariego Castiñeira, Ana Cerezales Calviño, Marta Contreras Sánchez, Ramón Fernández Varela, Santiago J. Freire Castro, Ana Padín Trigo, Rafael Prieto Jarel, Fátima Raad Varea, Laura Ramos Alonso, Francisco Javier Sanmartín Pensado, David Vieito Porto.

<u>H. Clinico San Carlos. Madrid</u>

Inés Armenteros Yeguas, Javier Azaña Gómez, Julia Barrado Cuchillo, Irene Burruezo López, Noemí Cabello Clotet, Alberto E. Calvo Elías, Elpidio Calvo Manuel, Carmen María Cano de Luque, Cynthia Chocron Benbunan, Laura Dans Vilan, Ester Emilia Dubon Peralta, Vicente Estrada Pérez, Santiago Fernandez, Marcos Oliver Fragiel Saavedra, José Luis García Klepzig, Maria del Rosario Iguarán, Esther Jaén Ferrer, Rubén Ángel Martín Sánchez, Manuel Méndez Bailón, Maria José Nuñez Orantos, Carolina Olmos Mata, Eva Orviz García, David Oteo Mata, Cristina Outon González, Juncal Perez-Somarriba, Pablo Pérez Mateos, Maria Esther Ramos Muñoz, Xabier Rivas Regaira, Iñigo Sagastagoitia Fornie, Alejandro Salinas Botrán, Miguel Suárez Robles, Maddalena Elena Urbano, Miguel Villar Martínez.

<u>H. Infanta Sofía. S. S. de los Reyes</u>

Rafael del Castillo Cantero, Rebeca Fuerte Martínez, Arturo Muñoz Blanco, José Francisco Pascual Pareja, Isabel Perales Fraile, Isabel Rábago Lorite, Llanos Soler Rangel, Inés Suárez García, Jose Luis Valle López.

<u>Hospital Universitario Dr. Peset. Valencia</u>

Juan Alberto Aguilera Ayllón, Arturo Artero Mora, María del Mar Carmona Martín, María José Fabiá Valls, Maria de Mar Fernández Garcés, Ana Belén Gómez Belda, Ian López Cruz, Manuel Madrazo López, Elisabet Mateo Sanchis, Jaume Micó Gandia, Laura Piles Roger, Adela Maria Pina Belmonte, Alba Viana García.

<u>Hospital Clínico de Santiago. Santiago de Compostela</u>

Maria del Carmen Beceiro Abad, Maria Aurora Freire Romero, Sonia Molinos Castro, Emilio Manuel Paez Guillan, María Pazo Nuñez, Paula Maria Pesqueira Fontan.

<u>Hospital Royo Villanova. Zaragoza</u>

Nicolás Alcalá Rivera, Anxela Crestelo Vieitez, Esther del Corral, Jesús Díez Manglano, Isabel Fiteni Mera, Maria del Mar Garcia Andreu, Martin Gerico Aseguinolaza, Claudia Josa Laorden, Raul Martinez Murgui, Marta Teresa Matía Sanz.

<u>H. U. Infanta Cristina. Parla</u>

Juan Miguel Antón Santos, Ana Belén Barbero Barrera, Coralia Bueno Muiño, Ruth Calderón Hernaiz, Irene Casado Lopez, José Manuel Casas Rojo, Andrés Cortés Troncoso, Mayte de Guzmán García-Monge, Francesco Deodati, Gonzalo García Casasola Sánchez, Elena Garcia Guijarro, Davide Luordo, María Mateos González, Jose A Melero Bermejo, Lorea Roteta García, Elena Sierra Gonzalo, Javier Villanueva Martínez.

<u>H. de Cabueñes. Gijón</u>

Ana María Álvarez Suárez, Carlos Delgado Vergés, Rosa Fernandez-Madera Martínez, Eva Fonseca Aizpuru, Alejandro Gómez Carrasco, Cristina Helguera Amezua, Juan Francisco López Caleya, María del Mar Martínez López, Aleida Martínez Zapico, Carmen Olabuenaga Iscar, María Luisa Taboada Martínez, Lara María Tamargo Chamorro.

<u>H. Santa Marina. Bilbao</u>

Maria Areses Manrique, Ainara Coduras Erdozain, Ane Elbire Labirua-Iturburu Ruiz.

<u>Hospital de Urduliz Alfredo Espinosa. Urdúliz</u>

María Aparicio López, Asier Aranguren Arostegui, Paula Arriola Martínez, Gorka Arroita Gonzalez, M^<u>a</u>^ Soledad Azcona Losada, Miriam García Gómez, Eduardo Garcia Lopez, Amalur Iza Jiménez, Alazne Lartategi Iraurgi, Esther Martinez Becerro, Itziar Oriñuela González, Isabel María Portales Fernández, Pablo Ramirez Sánchez, Beatriz Ruiz Estévez, Cristian Vidal Núñez.

<u>Hospital HLA Moncloa. Madrid</u>

Guillermo Estrada, Teresa Garcia Delange, Isabel Jimenez Martinez, Carmen Martinez Cilleros, Nuria Parra Arribas.

<u>H. del Henares. Coslada</u>

Jesús Ballano Rodríguez-Solís, Luis Cabeza Osorio, María del Pilar Fidalgo Montero, M^<u>a</u>^ Isabel Fuentes Soriano, Erika Esperanza Lozano Rincon, Ana Martín Hermida, Jesus Martinez Carrilero, Jose Angel Pestaña Santiago, Manuel Sánchez Robledo, Patricia Sanz Rojas, Nahum Jacobo Torres Yebes, Vanessa Vento.

<u>H. Nuestra Señora del Prado. Talavera de la Reina</u>

Sonia Casallo Blanco, Jeffrey Oskar Magallanes Gamboa.

<u>H. U. Torrevieja. Torrevieja</u>

Julio César Blázquez Encinar.

<u>H. U. La Fe. Valencia</u>

Dafne Cabañero, María Calabuig Ballester, Pascual Císcar Fernández, Ricardo Gil Sánchez, Marta Jiménez Escrig, Cristina Marín Amela, Laura Parra Gómez, Carlos Puig Navarro, José Antonio Todolí Parra.

<u>H. San Pedro. Logroño</u>

Diana Alegre González, Irene Ariño Pérez de Zabalza, Sergio Arnedo Hernández, Jorge Collado Sáenz, Beatriz Dendariena, Marta Gómez del Mazo, Iratxe Martínez de Narvajas Urra, Sara Martínez Hernández, Estela Menendez Fernández, Jose Luís Peña Somovilla, Elisa Rabadán Pejenaute.

<u>Hospital Universitario Ntra Sra Candelaria. Santa Cruz de Tenerife</u>

Lucy Abella, Andrea Afonso Díaz, Selena Gala Aguilera Garcia, Marta Bethencourt Feria, Eduardo Mauricio Calderón Ledezma, Sara Castaño Perez, Guillermo Castro Gainett, José Manuel del Arco Delgado, Joaquín Delgado Casamayor, Diego Garcia Silvera, Alba Gómez Hidalgo, Marcelino Hayek Peraza, Carolina Hernández Carballo, Rubén Hernández Luis, Francisco Javier Herrera Herrera, Maria del Mar Lopez Gamez, Julia Marfil Daza, María José Monedero Prieto, María Blanca Monereo Muñoz, María de la Luz Padilla Salazar, Daniel Rodríguez Díaz, Alicia Tejera, Laura Torres Hernández.

<u>H. U. San Juan de Alicante. San Juan de Alicante</u>

David Balaz, David Bonet Tur, Carles García Cervera, David Francisco García Núñez, Vicente Giner Galvañ, Angie Gómez Uranga, Javier Guzmán Martínez, Isidro Hernández Isasi, Lourdes Lajara Villar, Juan Manuel Núñez Cruz, Sergio Palacios Fernández, Juan Jirge Peris García, Andrea Riaño Pérez, José Miguel Seguí Ripoll, Philip Wikman-Jorgensen.

<u>H. U. San Agustin. Avilés</u>

Andrea Álvarez García, Víctor Arenas García, Alba Barragán Mateos, Demelsa Blanco Suárez, María Caño Rubia, Jaime Casal Álvarez, David Castrodá Copa, José Ferreiro Celeiro, Natalia García Arenas, Raquel García Noriega, Joaquin Llorente García, Irene Maderuelo Riesco, Paula Martinez Garcia, Maria Jose Menendez Calderon, Diego Eduardo Olivo Aguilar, Marta Nataya Solís Marquínez, Luis Trapiella Martínez, Andrés Astur Treceño García, Juan Valdés Bécares.

<u>H. U. Son Llàtzer. Palma de Mallorca</u>

Andrés de la Peña Fernández, Almudena Hernández Milián.

<u>H. de Mataró. Mataró</u>

Raquel Aranega González, Ramon Boixeda, Carlos Lopera Mármol, Marta Parra Navarro, Ainhoa Rex Guzmán, Aleix Serrallonga Fustier.

<u>H. Juan Ramón Jiménez. Huelva</u>

Francisco Javier Bejarano Luque, Francisco Javier Carrasco-Sánchez, Mercedes de Sousa Baena, Jaime Díaz Leal, Aurora Espinar Rubio, Maria Franco Huertas, Juan Antonio García Bravo, Andrés Gonzalez Macías, Encarnación Gutiérrez Jiménez, Constantino Lozano Quintero, Carmen Mancilla Reguera, Francisco Javier Martínez Marcos, Francisco Muñoz Beamud, Maria Perez Aguilera, Alícia Perez Jiménez, Virginia Rodríguez Castaño, Alvaro Sánchez de Alcazar del Río, Leire Toscano Ruiz.

<u>H. U. Reina Sofía. Córdoba</u>

Antonio Pablo Arenas de Larriva, Pilar Calero Espinal, Javier Delgado Lista, María Jesús Gómez Vázquez, Jose Jiménez Torres, Laura Martín Piedra, Javier Pascual Vinagre, María Elena Revelles Vílchez, Juan Luis Romero Cabrera, José David Torres Peña.

<u>H. Moisès Broggi. Sant Joan Despí</u>

Jose Loureiro Amigo, Melani Pestaña Fernández, Nicolas Rhyman, Nuria Vázquez Piqueras.

<u>H. U. Virgen de las Nieves. Granada</u>

Pablo Conde Baena, Joaquin Escobar Sevilla, Laura Gallo Padilla, Patricia Gómez Ronquillo, Pablo González Bustos, María Navío Botías, Jessica Ramírez Taboada, Mar Rivero Rodrígez.

<u>H. San Juan de la Cruz. Úbeda</u>

Marcos Guzmán Garcia, Francisco Javier Vicente Hernández.

<u>Hospital Costa del Sol. Marbella</u>

Victoria Augustín Bandera, María Dolores Martín Escalante.

<u>Complejo Asistencial Universitario de León. León</u>

Rosario Maria García Die, Manuel Martin Regidor, Angel Luis Martínez Gonzalez, Alberto Muela Molinero, Raquel Rodríguez Díez, Beatriz Vicente Montes.

<u>Hospital Clinic Barcelona. Barcelona</u>

Júlia Calvo Jiménez, Aina Capdevila Reniu, Irene Carbonell De Boulle, Emmanuel Coloma Bazán, Joaquim Fernández Solà, Cristina Gabara Xancó, Joan Ribot Grabalosa, Olga Rodríguez Núñez.

<u>Hospital Marina Baixa. Villajoyosa</u>

Javier Ena, Jose Enrique Gómez Segado.

<u>C. H. U. de Ferrol. Ferrol</u>

Hortensia Alvarez Diaz, Tamara Dalama Lopez, Estefania Martul Pego, Carmen Mella Pérez, Ana Pazos Ferro, Sabela Sánchez Trigo, Dolores Suarez Sambade, Maria Trigas Ferrin, Maria del Carmen Vázquez Friol, Laura Vilariño Maneiro.

<u>Hospital del Tajo. Aranjuez</u>

Ruth Gonzalez Ferrer, Raquel Monsalvo Arroyo.

<u>H. U. Marqués de Valdecilla. Santander</u>

Marta Fernández-Ayala Novo, José Javier Napal Lecumberri, Nuria Puente Ruiz, Jose Riancho, Isabel Sampedro Garcia.

<u>Hospital Torrecárdenas. Almería</u>

Luis Felipe Díez García, Iris El Attar Acedo, Bárbara Hernandez Sierra, Carmen Mar Sánchez Cano.

<u>Hospital Infanta Margarita. Cabra</u>

María Esther Guisado Espartero, Lorena Montero Rivas, Maria de la Sierra Navas Alcántara, Raimundo Tirado-Miranda.

<u>H. U. Severo Ochoa. Leganés</u>

Yolanda Casillas Viera, Lucía Cayuela Rodríguez, Carmen de Juan Alvarez, Gema Flox Benitez, Laura García Escudero, Juan Martin Torres, Patricia Moreira Escriche, Susana Plaza Canteli, M Carmen Romero Pérez.

<u>Hospital Insular de Gran Canaria. Las Palmas G. C</u>

Carlos Jorge Ripper.

<u>Hospital Valle del Nalón. Riaño (Langreo)</u>

Sara Fuente Cosío, César Manuel Gallo Álvaro, Julia Lobo García, Antía Pérez Piñeiro.

<u>H. U. del Vinalopó. Elche</u>

Francisco Amorós Martínez, Erika Ascuña Vásquez, Jose Carlos Escribano Stablé, Adriana Hernández Belmonte, Ana Maestre Peiró, Raquel Martínez Goñi, M. Carmen Pacheco Castellanos, Bernardino Soldan Belda, David Vicente Navarro.

<u>Hospital Alto Guadalquivir. Andújar</u>

Begoña Cortés Rodríguez.

<u>H. Francesc de Borja. Gandia</u>

Alba Camarena Molina, Simona Cioaia, Anna Ferrer Santolalia, José María Frutos Pérez, Eva Gil Tomás, Leyre Jorquer Vidal, Marina Llopis Sanchis, Mari Ángeles Martínez Pascual, Alvaro Navarro Batet, Mari Amparo Perea Ribis, Ricardo Peris Sanchez, José Manuel Querol Ribelles, Silvia Rodriguez Mercadal, Ana Ventura Esteve.

<u>H. G. U. de Castellón. Castellón de la Plana</u>

Jorge Andrés Soler, Marián Bennasar Remolar, Alejandro Cardenal Álvarez, Daniela Díaz Carlotti, María José Esteve Gimeno, Sergio Fabra Juana, Paula García López, María Teresa Guinot Soler, Daniela Palomo de la Sota, Guillem Pascual Castellanos, Ignacio Pérez Catalán, Celia Roig Martí, Paula Rubert Monzó, Javier Ruiz Padilla, Nuria Tornador Gaya, Jorge Usó Blasco.

<u>H. Santa Bárbara. Soria</u>

Marta Leon Tellez.

<u>C. A. U. de Salamanca. Salamanca</u>

Gloria María Alonso Claudio, Víctor Barreales Rodríguez, Cristina Carbonell Muñoz, Adela Carpio Pérez, María Victoria Coral Orbes, Daniel Encinas Sánchez, Sandra Inés Revuelta, Miguel Marcos Martín, José Ignacio Martín González, José Ángel Martín Oterino, Leticia Moralejo Alonso, Sonia Peña Balbuena, María Luisa Pérez García, Ana Ramon Prados, Beatriz Rodríguez-Alonso, Ángela Romero Alegría, Maria Sanchez Ledesma, Rosa Juana Tejera Pérez.

<u>H. U. de Canarias. Santa Cruz de Tenerife</u>

Julio Cesar Alvisa Negrin, José Fernando Armas González, Lourdes González Navarrete, Iballa Jiménez, María Candelaria Martín González, Miguel Nicolas Navarrete Lorite, Paula Ortega Toledo, Onán Pérez Hernández, Alina Pérez Ramírez.

<u>H. de Poniente. Almería</u>

Juan Antonio Montes Romero, Encarna Sánchez Martín, Jose Luis Serrano Carrillo de Albornoz, Manuel Jesus Soriano Pérez.

<u>H. U. Lucus Augusti. Lugo</u>

Raquel Gómez Méndez, Ana Rodríguez Álvarez.

<u>H. San Pedro de Alcántara. Cáceres</u>

Angela Agea Garcia, Javier Galán González, Luis Gámez Salazar, Eva Garcia Sardon, Antonio González Nieto, Itziar Montero Días, Selene Núñez Gaspar, Alvaro Santaella Gomez.

<u>H. U. del Sureste. Arganda del Rey</u>

Jon Cabrejas Ugartondo, Ana Belén Mancebo Plaza, Arturo Noguerado Asensio, Bethania Pérez Alves, Natalia Vicente López.

<u>H. de Pozoblanco. Pozoblanco</u>

José Nicolás Alcalá Pedrajas, Antonia Márquez García, Inés Vargas.

<u>Hospital Doctor José Molina Orosa. Arrecife (Lanzarote)</u>

Virginia Herrero García, Berta Román Bernal.

<u>H. Nuestra Señora de Sonsoles. Ávila</u>

Alaaeldeen Abdelhady Kishta.

<u>C. H. U. de Badajoz. Badajoz</u>

Rafael Aragon Lara, Inmaculada Cimadevilla Fernandez, Juan Carlos Cira García, Gema Maria García García, Julia Gonzalez Granados, Beatriz Guerrero Sánchez, Francisco Javier Monreal Periáñez, Maria Josefa Pascual Perez.

<u>H. G. U. de Elda. Elda</u>

Carmen Cortés Saavedra, Jennifer Fernández Gómez, Borja González López, María Soledad Hernández Garrido, Ana Isabel López Amorós, Maria de los Reyes Pascual Pérez, Andrea Torregrosa García.

<u>H. U. Puerta del Mar. Cádiz</u>

José Antonio Girón González, Susana Fabiola Pascual Perez, Cristina Rodríguez Fernández-Viagas, Maria José Soto Cardenas.

<u>Hospìtal de Montilla. Montilla</u>

Ana Cristina Delgado Zamorano, Beatriz Gómez Marín, Adrián Montaño Martínez, Jose Luis Zambrana García.

<u>H. Virgen de los Lirios. Alcoy (Alicante)</u>

M^<u>a</u>^ José Esteban Giner.

<u>H. Infanta Elena. Huelva</u>

María Gloria Rojano Rivero.

<u>H. de la Axarquía. Vélez-Málaga</u>

Antonio Lopez Ruiz.

<u>H. Virgen del Mar. Madrid</u>

Maria Jesus Gonzalez Juarez.

<u>Hospital do Salnes. Vilagarcía de Arousa</u>

Vanesa Alende Castro, Ana María Baz Lomba, Ruth Brea Aparicio, Marta Fernandez Morales, Jesus Manuel Fernandez Villar, Maria Teresa Lopez Monteagudo, Cristina Pérez García, Lorena María Rodríguez Ferreira, Maria Begoña Valle Feijoo.

## REFERENCES

1. World Health Organization. WHO coronavirus disease (COVID-19) dashboard. Available from: https://covid19.who.int/?gclid=EAIaIQobChMIjcGK1NDe6QIVWojVCh0vhQLLEAAYASABEgKs2fD_BwE. Last accessed: October, 30, 2020.

2. Venogupal U, Jilani N, Rabah S, Shariff MA, Jawed M, Medez Batres A, et al. SARS-CoV-2 seroprevalence among health care workers in a New York city hospital: a cross-sectional analysis during the COVID-19 pandemic. Int J Infect Dis 2020; http://doi.org/10.1016/j.intid.2020.10.036.

3. Rudberg AS, Havervall S, Manberg A, Falk AJ, Aguilera K, Ng, H, et al. SARS-CoV-2 exposure, symptoms and seroprevalence in healthcare workers in Sweden. Nat Commun 2020; 11: 5064.

4. Herzberg J, Vollmer T, Fischer B, Becher H, Becker AK, Sahly A, et al. A prospective sero-epidemiological evaluation of SARS-CoV-2 among health care workers in a German secondary care hospital. Int J Infect Dis 2020; http://doi.org/10.1016/j.ijid.2020.10.026

5. Sahu AK, Amrithanand VT, Mathew R, Aggarwal P, Nayer J, Bhoi S. COVID-19 in health care workers – A systematic review and meta-analysis. Am J Emerg Med 2020; 38: 1727–31.

6. España: Ministerio de Sanidad. Situación de COVID-19 en España. [Internet]. Centro de Coordinación de Alertas y Emergencias Sanitarias. Enfermedad por el coronavirus (COVID-19). Available from: https://covid19.isciii.es/. Last accesed: May 24, 2020.

7. Shah ASV, Wood R, Gribben C, Caldwell C, Bishop J, Wier M, et al. Risk of hospital admission with coronavirus disease 2019 in healthcare workers and theirs households: nationwide linkage cohort study. BMJ 2020; 371: m3582.

8. Hughes MM, Groenewold MR, Lessem SE, Xu K, Usseru EN, Wiegand RE, et al. Update: Characteristics of health care personnel with COVID-19 — United States, February 12–July 16, 2020. MMWR 2020; 69: 1364–8.

9. Lai X, Wang M, Qin C, Tan L, Ran L, Chen D, et al. Coronavirus disease 2019 (COVID-19) infection among health care workers and implications for prevention measures in a tertiary hospital in Wuhan, China. JAMA Network Open 2020; 3: e209666.

10. Saraiva Duarte MM, Claudino Haslett MI, Alves de Freitas LJ, Nulle Gomes MT, Castanha da Silva DC, Percio J, et al. Description of COVID-19 hospitalized health care worker cases in the first nine weeks of the pandemic, Brazil, 2020. Epidemiol Serv Saude 2020; 29: e20200277.

11. Wang X, et al. Clinical characteristics of 80 hospitalized frontline medical workers infected with COVID-19 in Wuhan, China. J Hosp Inf 2020; 105: 399–403.

12. Dominguez-Varela IA. High mortality among health personnel with COVID-19 in Mexico. Disaster Med Public Health Prep 2020; Oct 13: 1–4.

13. Nienhaus A, Hod R. COVID-19 among health workers in Germany and Malaysia. Int J Environ Res Public Health 2020; 17: 4881.

14. Casas Rojo JM, Antón Santos JM, Nuñez-Cortés JM, Lumbreras C, Ramos Rincón JM, Roy-Vallejo E, et al; for the SEMI-COVID-19 Network. Clinical characteristics of patients hospitalized with COVID-19 in Spain: results from the SEMI-COVID-19 Network. Rev Clin Esp 2020; 220: 480–94.

15. Rodilla E, Saura A, Jiménez I, Mendizábal A, Pineda-Cantero A, Lorenzo-Hernández, et al. Association of hypertension with all-cause mortality among hospitalized patients with COVID-19. J Clin Med 2020; 9: E3136.

16. Ramos Rincón JM, Buonaiuto V, Ricci M, Martín-Carmona J, Paredes-Ruiz D, Calderón-Moreno M, et al; for the SEMI-COVID-19 Network. Clinical characteristics and risk factors for mortality in very old patients hospitalized with COVID-19 in Spain. J Gerontol A Biol Sci Med Sci 2020; Oct 26: glaa243.

17. Charlson ME, Pompei P, Ales KL, MacKenzie CR. A new method of classifying prognostic comorbidity in longitudinal studies: development and validation. J Chron Dis 187; 40: 373–83.

18. Guan W, Ni Z, Hu Y, Liang W, Ou C, He J, et al. Clinical Characteristics of Coronavirus Disease 2019 in China. N Engl J Med 2020; 382: 1708–20.

19. Richardson S, Hirsch JS, Narasimhan M, Crawford JM, McGinn T, Davidson KW, et al. Presenting Characteristics, Comorbidities, and Outcomes Among 5700 Patients Hospitalized With COVID-19 in the New York City Area. JAMA 2020; e206775.

20. Vandercam G, Simon A, Scohy A, Belkhir L, Kabamba B, Rodriguez Villalobos H, et al. Clinical characteristics and humoral immune response in healthcare workers with COVID-19 in a teaching hospital in Belgium. J Hosp Infection 2020; 106: 713–20.

21. Antonio-Villa NE, Bello-Chavolla OY, Vargas-Vázquez A, Fermín-Martínez CA, Márquez-Salinas A, Bahena-López JP. Health-care workers with COVID-19 living in Mexico City: clinical characterization and related outcomes. Clin Infect Dis 2020; Sep 28, ciaa1487. doi: 10.1093/cid/ciaa1487.

22. Last J. A dictionary of epidemiology. 3rd ed. Oxford, UK: Oxford University Press; 1995.

23. Ho FK, Petermann-Rocha F, Gray SR, Jani BD, Katikireddi SV, Niedzwiedz CL, et al. Is older age associated with COVID-19 mortality in the absence of other risk factors? General population cohort study of 470,034 participants. PLoS ONE 2020; 15: e0241824.

24. Zheng Z, Peng F, Hu B, Zhao J, Liu H, Peng J, et al. Risk factors of critical & mortal COVID-19 cases: A systematic literature review and meta-analysis. J Infect 2020; 81: e16–25.

25. Yang J, Zheng Y, Gou X, Pu K, Chen Z, Guo Q, et al. Prevalence of comorbidities and its effects in coronavirus disease 2019 patients: A systematic review and meta-analysis. Int J Infect Dis 2020; 94:91–5.

26. Zhou Y, Yang Q, Chi J, Dong B, Lv W, Shen L, et al. Comorbidities and the risk of severe or fatal outcomes associated with coronavirus disease 2019: A systematic review and meta–analysis. Int J Infect Dis 2020; 99: 47–56.

